# Comparing the value of left atrial strain and HFA-PEFF score in diagnosing heart failure with preserved ejection fraction: a cross-sectional study

**DOI:** 10.1101/2024.05.16.24307480

**Authors:** Hai Nguyen Ngoc Dang, Thang Viet Luong, Bang Hai Ho, Tien Anh Hoang, Anh Xuan Mai, Hung Minh Nguyen

**Author notes:** Corresponding Author: Tien Anh Hoang, University of Medicine and Pharmacy, Hue University, 06 Le Loi Street, Hue city, Thua Thien Hue, 530000, Vietnam. These authors contributed equally to this work (Hai Nguyen Ngoc Dang, Thang Viet Luong).

## Abstract

**Objectives:** Heart failure with preserved ejection fraction (HFpEF) has a high hospitalization rate. While recent guidelines recommend specific parameters like E/e’, e’ velocity, and left atrial volume index (LAVI) for diagnosing HFpEF, their clinical accuracy remains limited. Left atrial (LA) strain has emerged as a potential diagnostic parameter, yet its role in the Vietnamese population is unclear. This study aims to evaluate LA strain’s diagnostic role in HFpEF among Vietnamese patients, exploring its relationship with established parameters of left ventricle (LV) diastolic function to determine its potential utility as a diagnostic tool.

**Methods:** A cross-sectional study was conducted from 15/04/2022 to 01/12/2023, involving 118 subjects, including 49 patients with HFpEF and 69 individuals without cardiac dysfunction. The study subjects were evaluated for LA strain and HFA-PEFF score. Diagnostic criteria for HFpEF were based on the 2021 European Society of Cardiology guidelines for diagnosing and treating acute and chronic heart failure.

**Results:** LA strain including LA reservoir (LASr), conduit (LAScd), and contractile (LASct) functions, in the HFpEF group were 20.80% [26.50 - 13.30], 9.08 ± 6.18%, and 10.89 ± 5.16%, respectively. The control group had corresponding LASr, LAScd, and LASct values of 34.45% [38.07 - 31.14], 17.33 ± 5.72%, and 17.38 ± 4.41% (p < 0.001). The area under the curve (AUC) for LASr, LAScd, LAScr, HFA-PEFF score, LAVI, and GLS to diagnose HFpEF was 0.852, 0.770, 0.778, 0.890, 0.615, and 0.701, respectively. Comparing the AUCs for diagnosing HFpEF between LASr and HFA-PEFF score, no difference was found with p = 0.419.

**Conclusion:** LA strain has a diagnostic value equivalent to the HFA-PEFF score in diagnosing HFpEF. These indices could be incorporated into the existing guidelines to enhance the diagnosis of HFpEF.

**Strengths and limitations of this study:**

- The study provides valuable data specific to the Vietnamese population, enhancing the understanding of heart failure with preserved ejection fraction and potentially leading to more tailored diagnostic strategies and treatments.
- This study directly evaluates two diagnostic tools by comparing left atrial strain with the HFA-PEFF score, which can help clinicians choose the most appropriate method for diagnosing HFpEF in clinical practice.
- The inclusion of left atrial strain as a diagnostic metric is relatively novel. It may provide new insights into the pathophysiology of HFpEF, offering a potential alternative or complement to existing diagnostic criteria.
- The findings can have immediate clinical implications, potentially improving the accuracy of HFpEF diagnosis and leading to better patient outcomes through more precise treatment plans.
- Our study has limitations, such as a relatively small sample size, sole location, and technical constraints. Addressing these limitations through further research will enhance the robustness and applicability of the findings.

## Introduction

Heart failure with preserved ejection fraction (HFpEF) is defined as heart failure with an ejection fraction of 50% or higher at diagnosis, affecting approximately 32 million people worldwide. Patients with HFpEF are hospitalized about 1.4 times per year and have an annual mortality rate of around 15%.^1^ Left ventricular (LV) diastolic dysfunction plays a fundamental role, overarching in the pathophysiology of HFpEF.^2^

The recent recommendations of the American Society of Echocardiography/European Society of Cardiovascular Imaging in 2016 used parameters such as E/e’, septal e’ velocity, lateral e’ velocity, maximum tricuspid valve regurgitation flow (TRV) and left atrial volume index (LAVI) to evaluate LV diastolic dysfunction as well as to diagnose HFpEF.^3^ However, in clinical practice, the recommendations parameters have limited precision in diagnosing HFpEF.^4^ Assessing left atrial (LA) function has recently become critical in cardiac evaluation.^5^ The LA function encompasses three primary aspects: blood storage (reservoir function), blood conduction (conduit function), and ejection function (contractile function).^6^ Commonly used indices to assess LA function include LAVI and LA size. Increased LAVI is associated with prolonged chronic LV/LA pressure overload. However, LA size takes time to change, often significantly dilating in later stages, making LAVI less sensitive in the early stages.^7^

LA strain is a novel echocardiographic technique that provides a comprehensive evaluation of reservoir, conduit, and contractile functions. This method proves particularly valuable when changes are subtle and challenging to detect using conventional parameters such as LA dimensions and LAVI.^8^ While LA dimensions have been previously utilized, the role of LA function as a biomarker is increasingly under evaluation, both independently and in conjunction with LA size. LA strain serves as a tool to assess LA function and can be measured throughout the cardiac cycle, enabling a thorough and comprehensive evaluation of LA reservoir, conduit, and contractile functions.^7^ Additionally, LA strain offers the advantage of being a technique mostly independent of angle and less susceptible to influences from mitral annulus calcification and bundle branch block effects.^9^

Notably, impaired LA strain has been observed in HFpEF patients, indicating its potential diagnostic value.^4^ ^10^ ^11^ Studies conducted in the United Kingdom and China have demonstrated the utility of LA strain in assessing and diagnosing HFpEF.^9^ ^12^ However, its role in the Vietnamese population remains unexplored. Therefore, this study aims to evaluate the diagnostic role of LA strain in HFpEF among Vietnamese individuals, contributing to a deeper understanding of its applicability in clinical practice.

## Materials & Methods

### Study Population

The study was conducted by the Declaration of Helsinki, and approved by The Institutional Ethics Committee of Hue University of Medicine and Pharmacy (Approval number: H2022/034). From 15/04/2022 to 01/12/2023, we conducted a cross-sectional study that randomly selecting 1014 adults aged 18 and above, who visited the Hue University Hospital for medical examinations. The study participants were fully informed about the benefits of participating in the research, and they were only included in the study if they verbally consented during the interview. After exclusions, we included a total of 118 subjects in the data analysis, comprising 49 individuals diagnosed with HFpEF in the disease group, and 69 individuals without cardiac dysfunction in the control group. The sampling process is detailed in **Supplementary Figure 1**. Patients with HFpEF are evaluated according to the standards of ESC in 2021:^13^ (1) Symptoms of heart failure (pulmonary congestion or systemic congestion); (2) Normal LV ejection fraction LVEF ≥ 50%; (3) Objective evidence of structural and/or functional cardiac abnormalities consistent with LV diastolic dysfunction; (4) NT-proBNP ≥ 125 pg/mL. The four recommended variables for identifying diastolic dysfunction and their abnormal cutoff values are septal e′ < 7 cm/s or lateral e′ < 10 cm/s, average E/e′ ratio > 14, LAVI > 34 mL/m2, and TRV > 2.8 m/s. If more than half of the available parameters met these cutoff values, LV diastolic dysfunction was considered present. In cases where half of the parameters met the cutoff values (indeterminate), diastolic dysfunction was assumed to be present if patients had E/A > 2.0 or if ≥ 2 of the three parameters (E/e′ ratio, TRV, and LAVI) met the cutoff values.^14^ Exclusion criteria included patients who declined participation, severe valvular heart disease, heart failure with EF < 50%, and arrhythmias. Patients with unclear echocardiography images or images lacking clear visualization of the myocardial endocardial layer were also excluded from the study. The control group comprised 69 healthy adults undergoing health screening with no history of heart failure.

### Clinical data collection, laboratory tests, and transthoracic echocardiogram

The clinical data collected included personal and family medical histories and clinical variables obtained through direct interviews and medical records. N-Terminal pro-B-type natriuretic peptide (NT-proBNP) levels were measured using a Cobas 8000 analyzer.

The echocardiographic procedure followed the American Society of Echocardiography guidelines for performing a comprehensive transthoracic echocardiographic examination in adults. During the procedure, the machine recorded the electrocardiogram alongside the echocardiographic images during the echocardiography procedure. All echocardiograms included in the final data analysis were performed on patients with normal sinus rhythm.^15^

### Left atrial and left ventricular strain analysis

Echocardiography images in DICOM format, meeting acceptable image quality standards, were uploaded to Philips QLAB Cardiovascular ultrasound quantification software Cardiac Analysis version 15. We conducted LA strain assessment in both the two-chamber and four-chamber views, setting reference points at the onset of the P wave in the cardiac cycle. Measurements of LA strain were acquired during the reservoir, conduit, and contractile phases of LA function, designated as LA strain reservoir function (LASr), LA strain conduit function (LAScd), and LA strain contractile function (LASct), respectively. For LV strain analysis, endocardial borders were traced on the end-systolic frame in three apical views (4-chamber, 2-chamber, and 3-chamber), with end-systole defined by the QRS complex or as the smallest LV volume during the cardiac cycle. The software tracked speckles along the endocardial border and myocardium throughout the cardiac cycle, automatically computing peak longitudinal strain and generating regional data from six segments, as well as an average value for each view. For patients with sinus rhythm, analyses were performed on a single cardiac cycle; for those with atrial fibrillation, strain values were calculated as the average of three cardiac cycles.^16^ One strain specialist in the core laboratory, who was blinded to the patients’ other data, performed all strain measurements.

LV and LA strain results are conventionally represented as negative values. However, for convenience in analysis and display, we utilized the absolute values of these results. The detailed methodology is illustrated in **Supplementary Figure 2**.

### Calculation of the HFA-PEFF score

The Heart Failure Association-PEFF (HFA-PEFF) score comprises functional, morphological, and biomarker domains (**Supplementary Table 1**). A patient can score zero, minor (1 point), or major (2 points) for each domain, and then those subscores are summed to produce a total score that ranges from 0 to 6 points. The total score is classified as low likelihood (0 – 1 point), intermediate likelihood (2 – 4 points), and high likelihood (5 – 6 points).^17^

### Statistics

We performed all statistical analyses using SPSS Version 26 (IBM, New York, United States), MedCalc Software Version 22.019 (MedCalc Software, Ostend, Belgium), and GraphPad Prism Version 10 (GraphPad Software, Boston, United States). Continuous variables were presented as mean ± standard deviation for normally distributed variables, as determined by the Kolmogorov-Smirnov test. Non-normally distributed variables were expressed as median values with interquartile ranges (25th-75th percentile). Categorical variables were reported as frequencies and percentages. We assessed intergroup differences in categorical variables using Fisher’s exact test, while differences in continuous variables were analyzed using the unpaired T-test or the Mann-Whitney U test, as appropriate. To assess the correlations between echocardiographic indices, NT-proBNP, and HFA-PEFF score, we used Spearman’s correlation coefficient (r_s_). The area under the curve (AUC) was determined using the Wilson/Brown method to diagnose HFpEF. We conducted AUC comparisons to assess the diagnostic value of strain compared to existing guideline criteria, employing the DeLong method.^18^ We randomly selected ten subjects from the control group and ten from the disease group to evaluate the intraclass correlation coefficient (ICC). The intraobserver and interobserver variability of LASr, LAScd, and LASct were assessed using the ICC and coefficient of variation. For intraobserver variability, the same operator independently remeasured the data after a 2-week interval. A second operator, blinded to the initial measurements, reanalyzed the data for interobserver variability. All statistical tests were two-sided, and a P-value of < 0.05 was considered significant.

## Results

### Baseline characteristics

**Table 1** displays age, sex, BSA, and BMI between the control and disease group, showing no statistically significant differences. The NT-proBNP concentration in the HFpEF group exhibited a non-normal distribution, with a median of 663.20 pg/mL (Quartile: 286.55 pg/mL – 1417.00 pg/mL). Additionally, LA strain indices in the HFpEF group were lower than in the control group, with all differences being statistically significant. Further details are illustrated in **Table 1**.

**Table 1:**
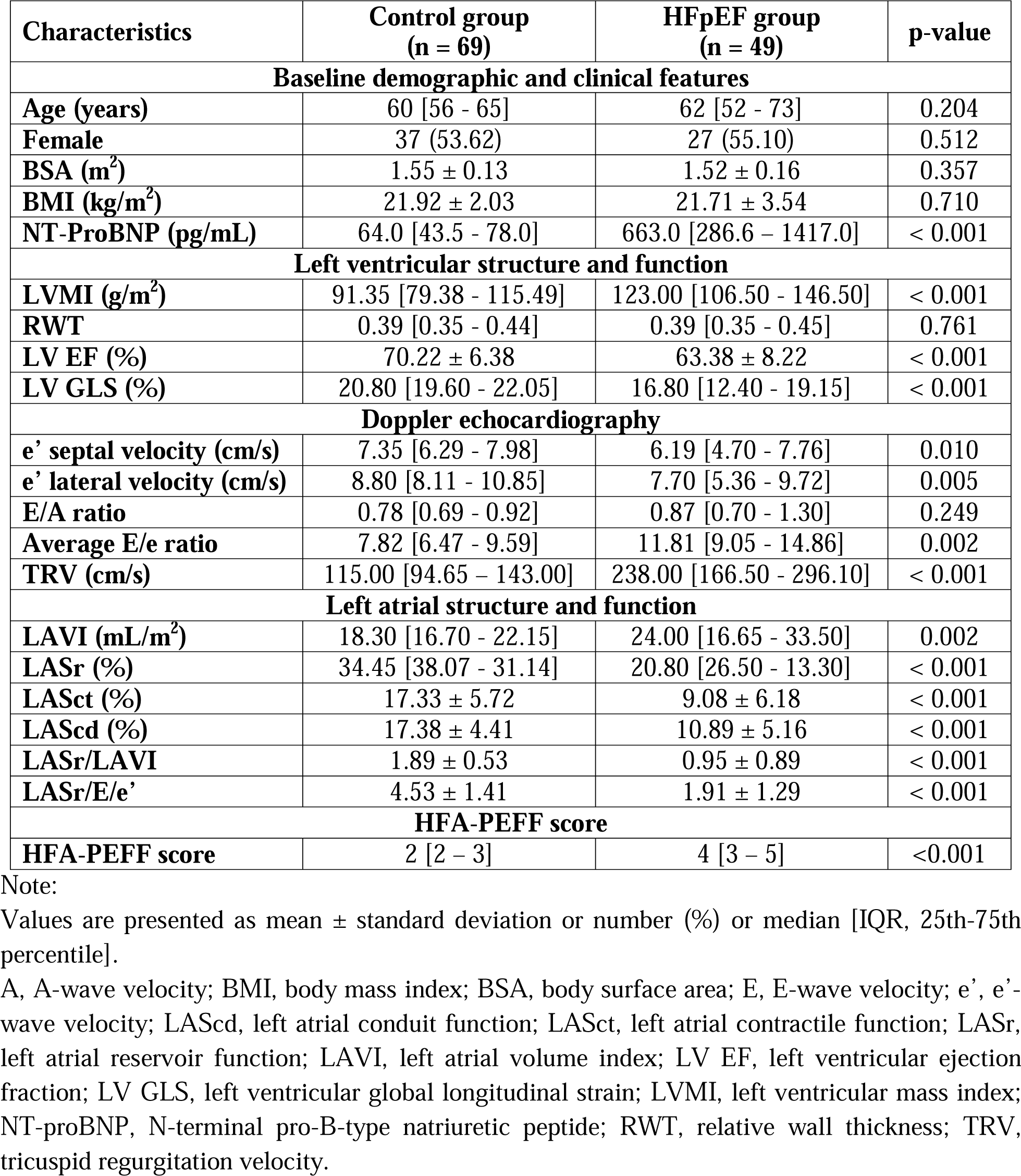
General characteristics of study subjects.

### Correlation analysis of the left atrial strains

**Figure 1** depicts the correlation between echocardiography indices, NT-proBNP, and HFA-PEFF score. LASr, LAScd, and LASct showed an inverse correlation with the HFA-PEFF score and NT-proBNP while also correlating with measured cardiac function indices. **Figure 1** demonstrates detailed parameters.

**Figure 1.**
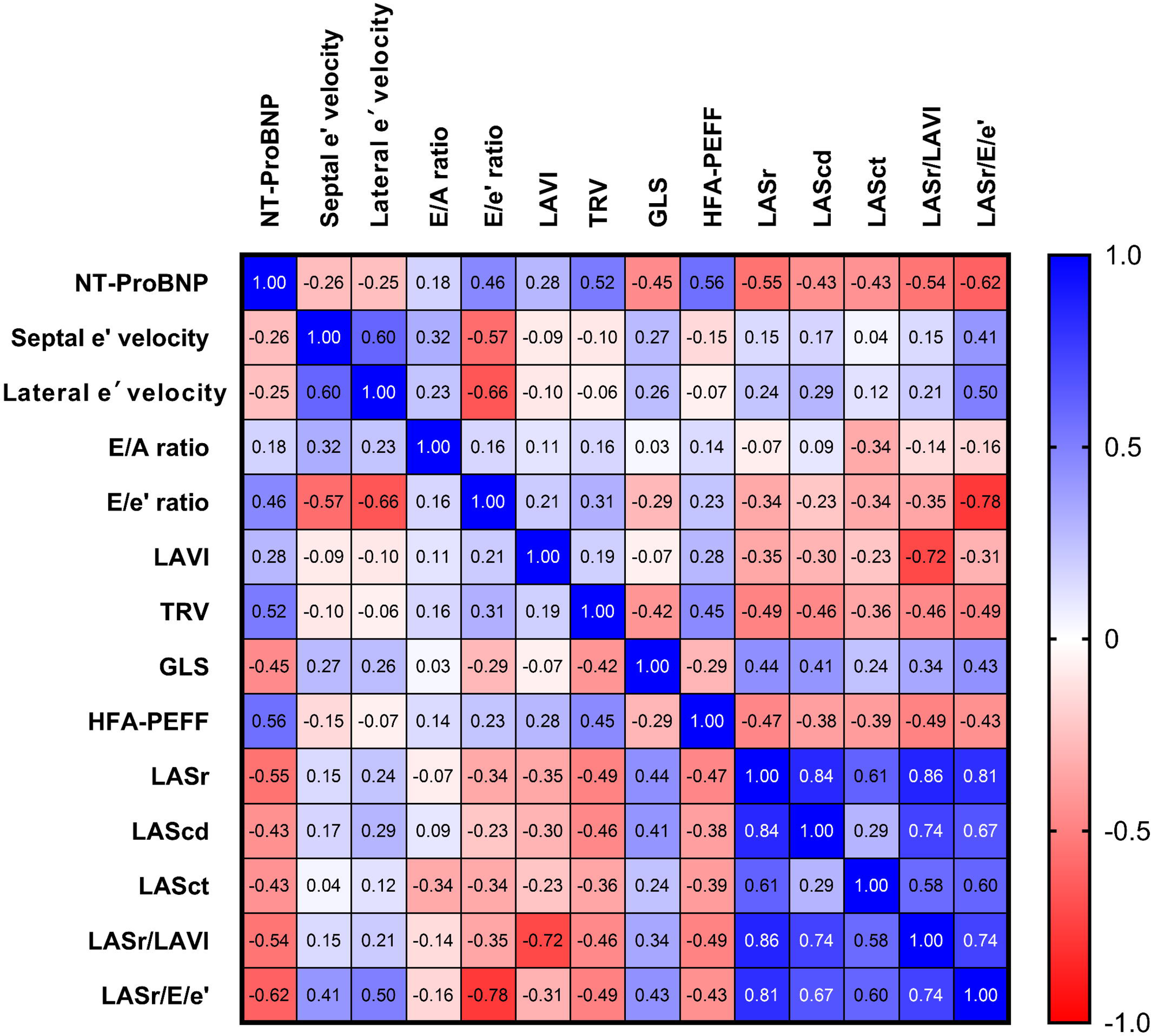
Heatmap depicts the correlation between echocardiography indices, NT-ProBNP, and HFA-PEFF. A: A-wave velocity; E: E-wave velocity; e’: e’-wave velocity; HFA-PEFF: Heart Failure Association-PEFF; LAScd: left atrial conduit function; LASct: left atrial contractile function; LASr: left atrial reservoir function; LAVI: left atrial volume index; LV GLS: left ventricular global longitudinal strain; NT-proBNP: N-terminal pro-B-type natriuretic peptide; TRV: tricuspid regurgitation velocity.

### The value of the left atrial strains and other echocardiographic parameters in diagnosing HFpEF

**Table 2** illustrates that the indices LASr (AUC = 0.852), LAScd (AUC = 0.770), LASct (AUC = 0.778), HFA-PEFF score (AUC = 0.890) exhibit high accuracy in diagnosing HFpEF. The cutoff points, sensitivity, and specificity of LASr are 29.85%, 83.67%, and 82.61%, respectively. Further detailed information is presented in **Table 2**.

**Table 2:**
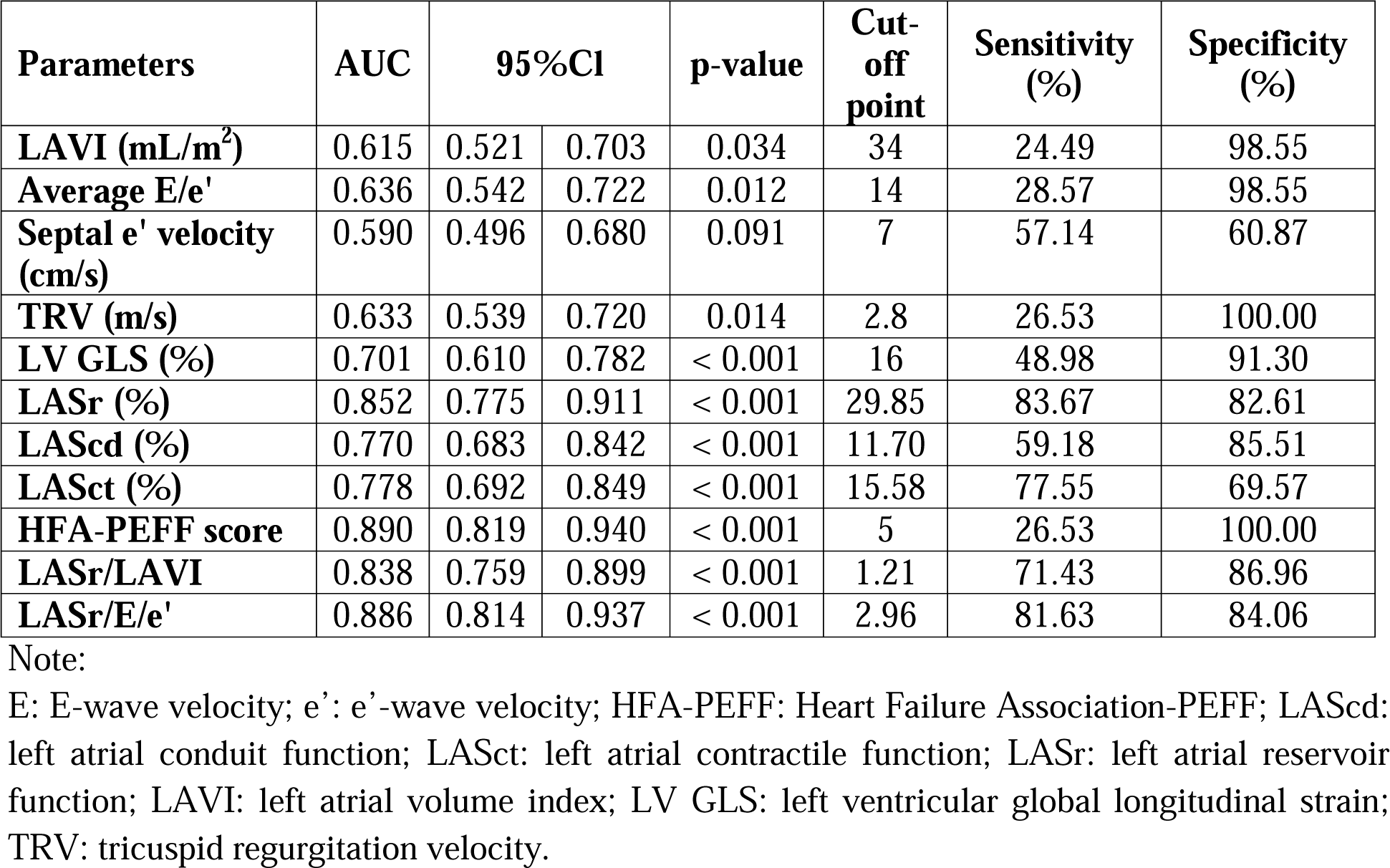
The performance of the LA strain parameters and existing criteria in diagnosing HFpEF.

**Table 3** reveals the area under the ROC curve for diagnosing HFpEF of HFA-PEFF score and LASr, showing no difference (p = 0.419). Moreover, the AUC of conventional echocardiographic indices (LAVI, E/e’, e’, TRV) with HFA-PEFF score and LASr differed significantly. Further detailed information is provided in **Table 3**.

**Table 3:**
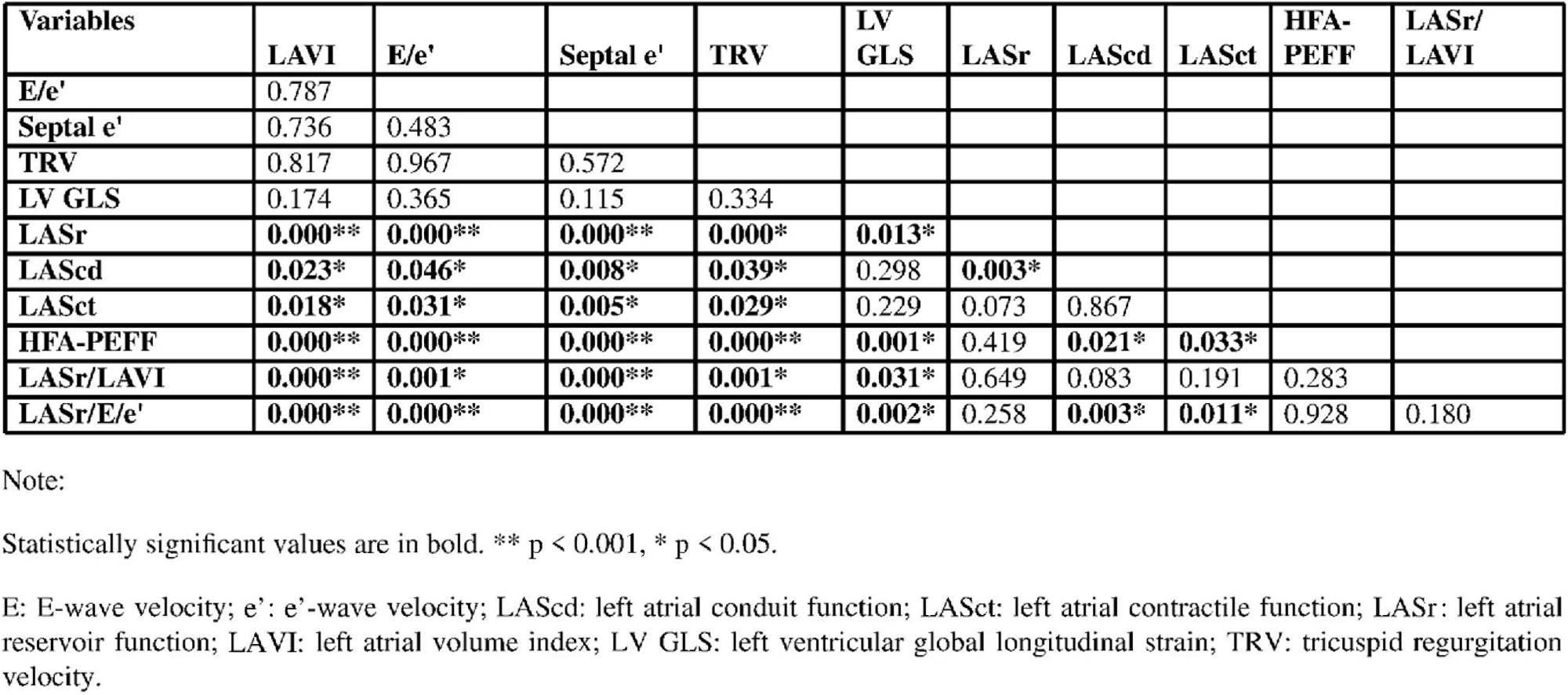
Correlation matrix with p values between AUC values of variables.

### Reliability of the left atrial strain measurements

**Figure 2** presents the intraobserver and interobserver variability for LA strain measurements. The parameters LASr, LAScd, and LASct demonstrated good reproducibility, indicated by high ICC values.

**Figure 2.**
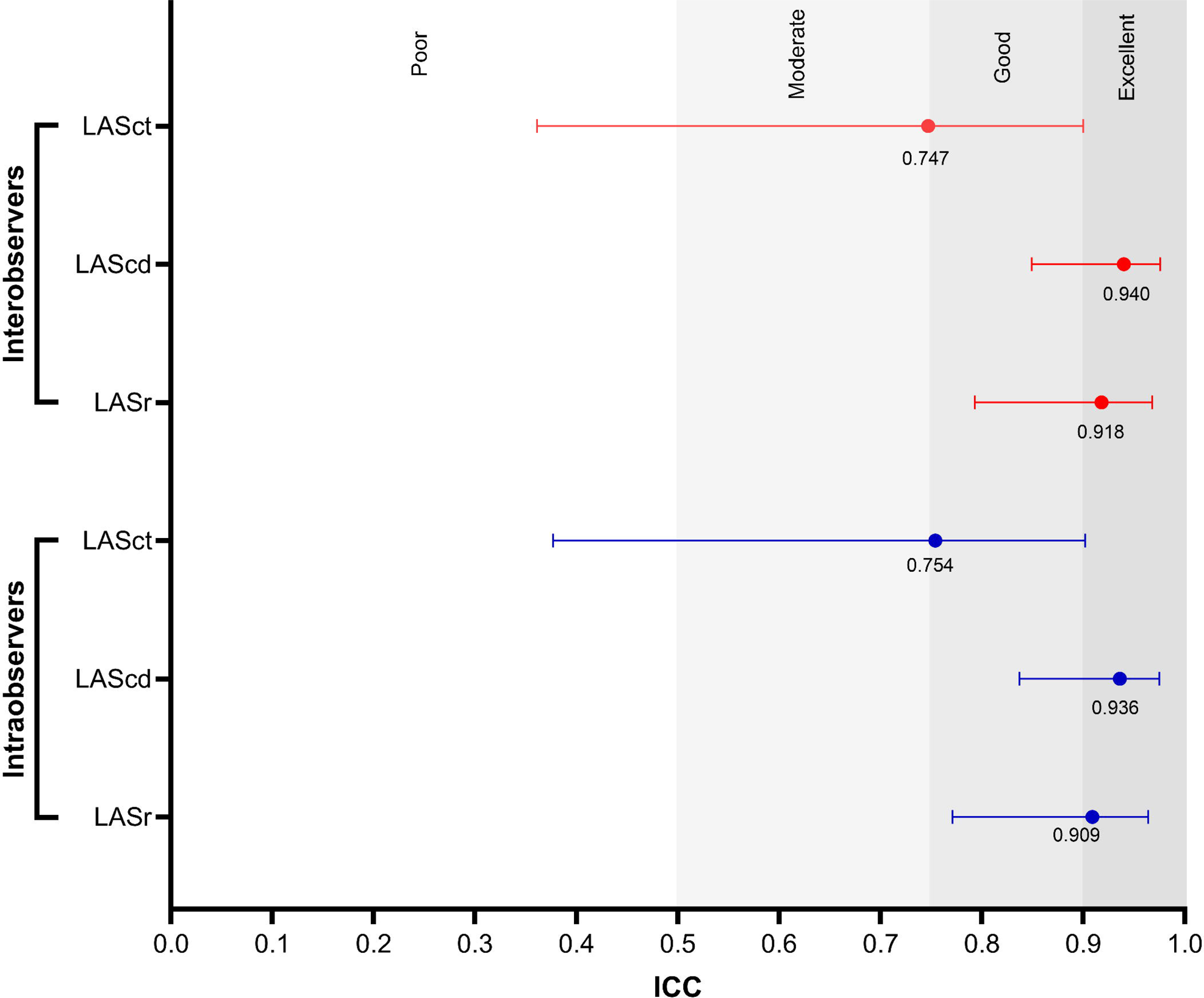
Reliability of LA strain measurements. LAScd: left atrial conduit function; LASct: left atrial contractile function; LASr: left atrial reservoir function; ICC: Intraclass correlation coefficient.

## Discussion

In this study, we evaluate the role of LA strain in diagnosing HFpEF. LA function indices such as LASr, LAScd, and LASct decreased significantly in the HFpEF group compared to the control group, with a p-value < 0.001. This finding is consistent with Aung et al.’s (2017) study on 83 patients, which also reported decreased LASr and LASct in the HFpEF group compared to the control group.^19^ Similar results were observed in several other studies where LASr, LAScd, and LASct were all reduced in HFpEF patients compared to the control group.^9^ ^12^ ^20^ ^21^ Therefore, our study confirms a significant decline in LA function in the HFpEF group compared to healthy subjects. Reddy et al. also highlighted the critical role of the LA in the progression of HFpEF. They suggested that LA strain reflects the overall LA function, which progressively deteriorates in chronic LV diastolic dysfunction, such as in patients with HFpEF.^9^

The American Echocardiography Association and the European Association of Cardiovascular Imaging (ASE/EACVI) recommended using echocardiography to diagnose HFpEF in patients with sinus rhythm using E/A ratio, LAVI, TRV, and E/e’.^3^ The ESC guidelines for the diagnosis of HFpEF are based on evidence of functional and structural alterations of the heart using the parameters E/e’, LVM, LAVI, septal e’ velocity, and lateral e’ velocity.^14^ However, these classic indicators still have many limitations, and many clinical practice cases encounter difficulties when the diagnosis falls into the “undetermined” state, when Doppler measurements cannot be made, such as tachycardia or severe mitral valve disease.^22^ Compared with Doppler echocardiography, the advantage of speckle tracking echocardiography when compared to conventional echocardiography is that it is independent of angle and less affected by mitral valve disease. On the other hand, the LA strain evaluates the function of the LA throughout the entire cardiac cycle rather than the functional state of a certain time point in the cardiac cycle.^4^ In our study, LA strain indices including LASr (AUC = 0.852), LAScd (AUC = 0.770), and LASct (AUC = 0.778) have high values in diagnosing HFpEF equivalent to the score HFA-PEFF score (AUC = 0.890). At the same time, the ability of LASr to diagnose HFpEF was superior to the LAVI (AUC = 0.615), E/e’ (AUC = 0.636), septal e’ velocity (AUC = 0.590), and TRV (AUC = 0.633) with p < 0.05. Many studies have also shown the superiority of LA strain indices in diagnosing HFpEF compared to commonly used classical indices.^9^ ^23^ ^24^

HFA-PEFF is a widely used scoring system for diagnosing HFpEF. However, evaluating this scoring system requires numerous parameters, including echocardiography, NT-proBNP, and atrial fibrillation diagnosis.^17^ ^25^ Our study shows that LA strain indices have demonstrated an AUC equivalent to the HFA-PEFF score. When comparing the AUC of LASr and HFA-PEFF score, our study found no significant difference in the HFpEF diagnostic value of LASr and HFA-PEFF score with p = 0.419. The 5-point HFA-PEFF score has 100% specificity, however, the sensitivity is low at only 26.53%, which can cause difficulties in applying this score in clinical practice.

In conclusion, LA parameters on speckle tracking echocardiography may be useful in diagnosing HFpEF. Integration into the 2016 EACVI/ASE criteria will improve diagnostic effectiveness with accuracy not inferior to conventional echocardiographic parameters.

### Limitations of the study

First, our study exclusively compares non-invasive indices for diagnosing HFpEF and refrains from using invasive interventions for evaluation or comparison with other invasive indices. We employed only one strain-analysis software platform and did not compare different software programs. Second, the speckle tracking echocardiography study was challenging due to image processing requirements, which led to the exclusion of many participants with incomplete data. This potential selection bias could impact the generalizability of our findings. Third, while our sample size for analysis is more significant that of some studies, it remains relatively small. More extensive studies are necessary to establish cutoff points relevant to clinical practice in Central Vietnam. Additionally, we conducted our study at a single location, which may limit the generalizability of our findings to other populations or settings. Variations in disease prevalence and characteristics across different populations or geographical locations could influence study outcomes. Fourth, during sample collection, technical limitations of Doppler echocardiography may have prevented us from obtaining all possible Doppler echocardiography indices for comparison with LA strain indices. Fifth, this study focused exclusively on subjects in sinus rhythm. However, atrial fibrillation represents a significant risk factor for HFpEF, necessitating further research to determine the optimal integration of LA strain parameters with conventional parameters for HFpEF diagnosis.

## Conclusions

The LA strain demonstrates diagnostic efficacy comparable to the HFA-PEFF score in diagnosing HFpEF. Integrating these indices into current guidelines could enhance future HFpEF diagnostics.

## Data Availability

Data cannot be shared publicly due to certain local ethical constraints. Researchers who meet the criteria for access to confidential data may contact the author, Hai Nguyen Ngoc Dang (via email at).

https://academic.oup.com/eurheartj/article/40/40/3297/5557740?login=false

## Acknowledgments

Not applicable.

## Author Contributions

Hai Nguyen Ngoc Dang: Conceptualization, Methodology, Acquisition, Investigation, Data curation, Writing – original draft, Writing – review & editing.

Thang Viet Luong: Conceptualization, Methodology, Acquisition, Investigation, Data curation, Writing – original draft, Writing – review & editing.

Bang Hai Ho: Conceptualization, Acquisition, Writing – original draft.

Tien Anh Hoang: Conceptualization, Acquisition, Writing – original draft.

Anh Xuan Mai: Conceptualization, Acquisition, Writing – original draft.

Hung Minh Nguyen: Conceptualization, Acquisition, Writing – original draft.

All authors have read and agreed to the published version of the manuscript.

---

**Supplementary Figure 1.**
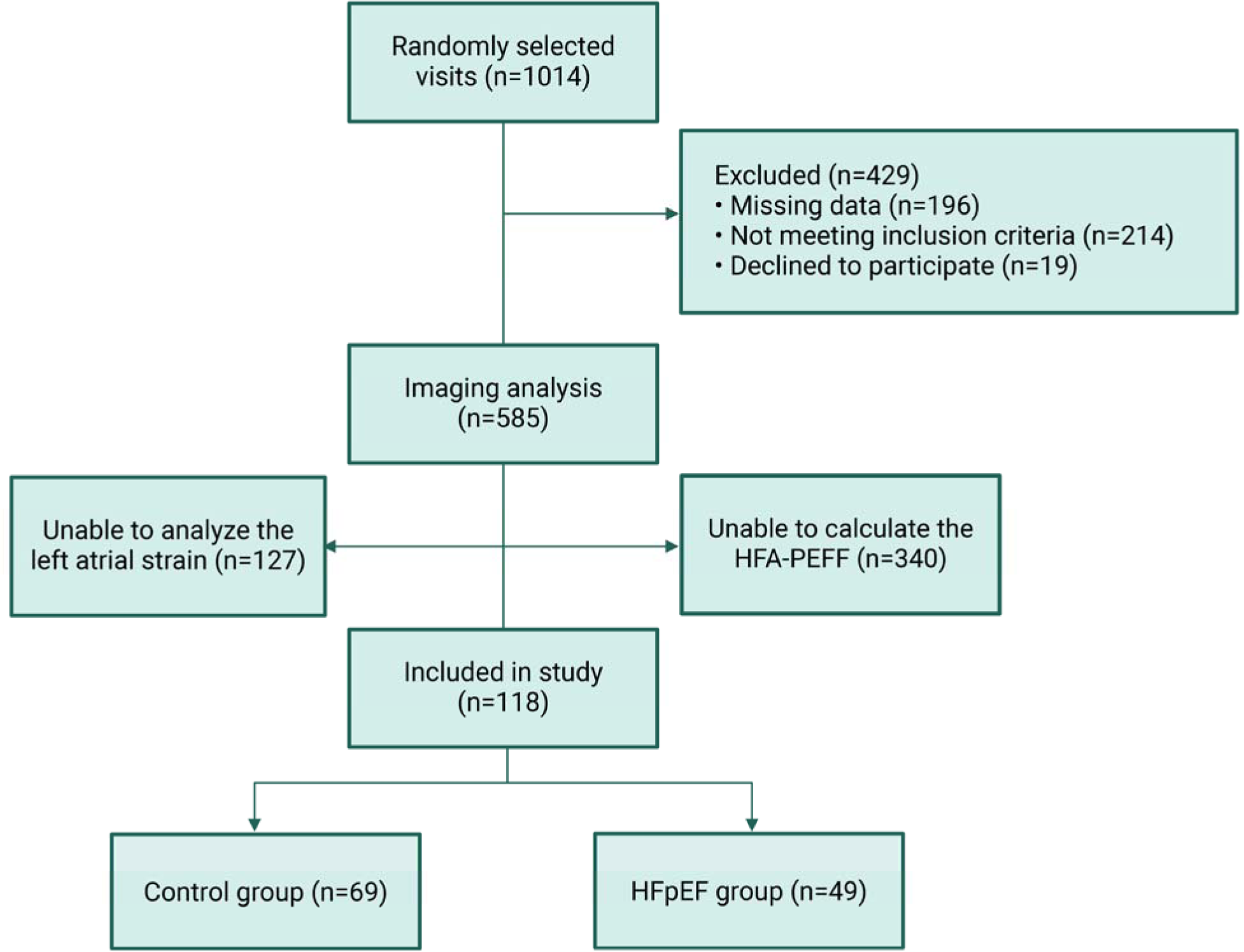
Flowchart illustrating the sample selection and exclusion process.

**Supplementary Figure 2.**
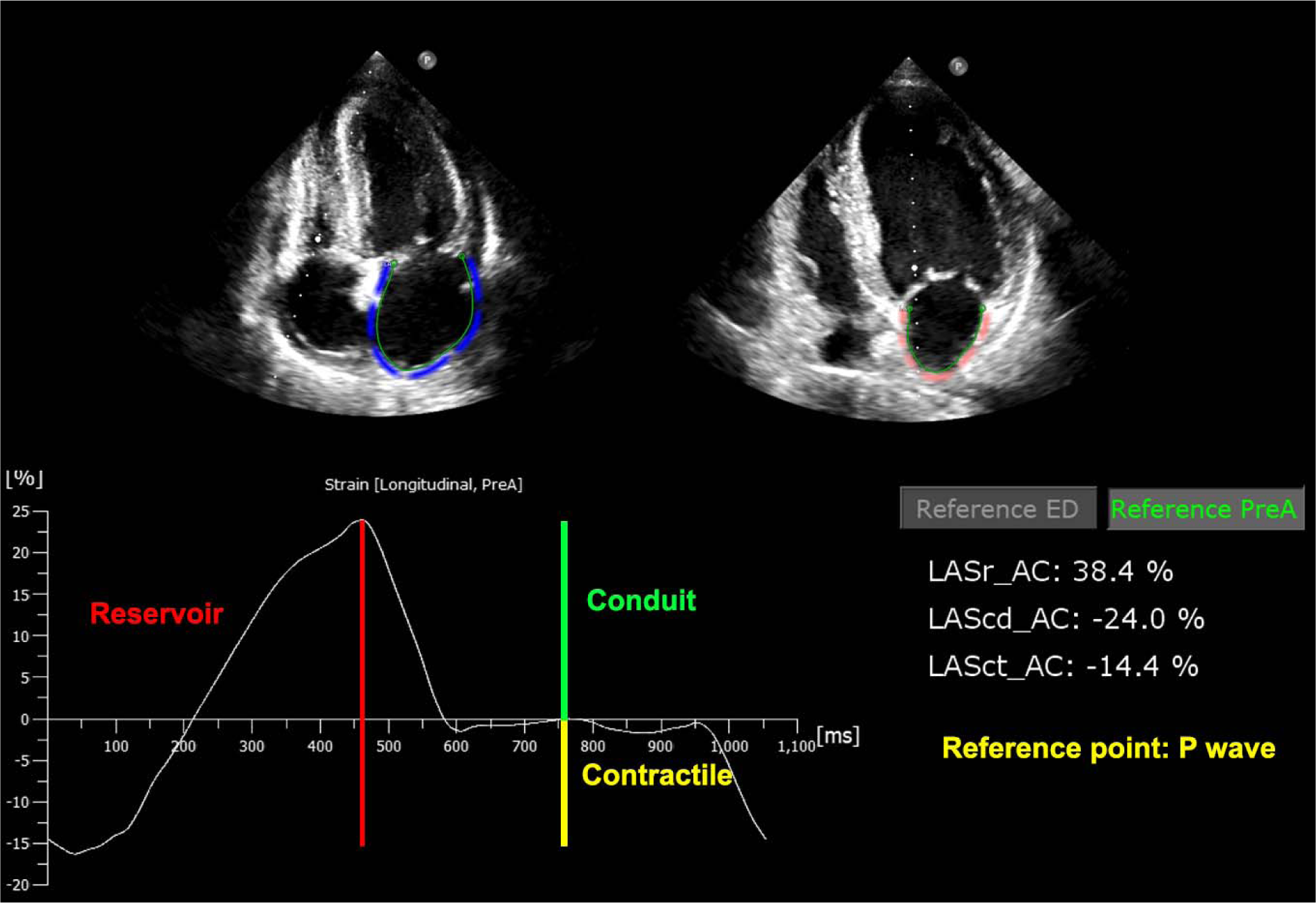
Parameters of LA strain on speckle tracking echocardiography. LASr (Reservoir) in red, LAScd (Conduit) in green, and LASct (Contractile) in yellow. LAScd: left atrial conduit function; LASct: left atrial contractile function; LASr: left atrial reservoir function.

**Supplementary Table 1.**
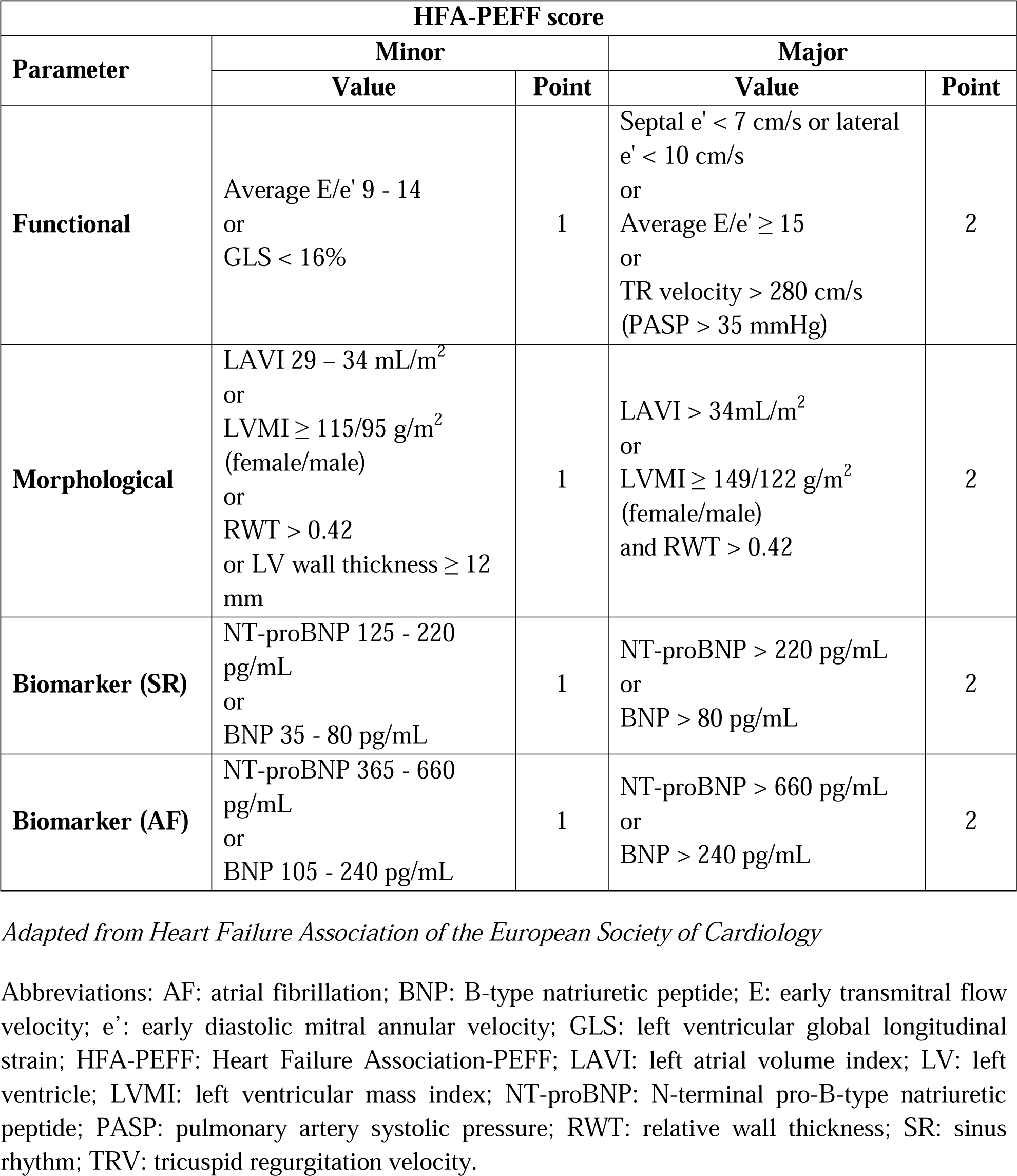
Calculation of HFA-PEFF score.

