## Supplementary figures and images for "Comparing the value of left atrial strain and HFA-PEFF score in diagnosing heart failure with preserved ejection fraction: a cross-sectional study"

### Supplementary Figure 1

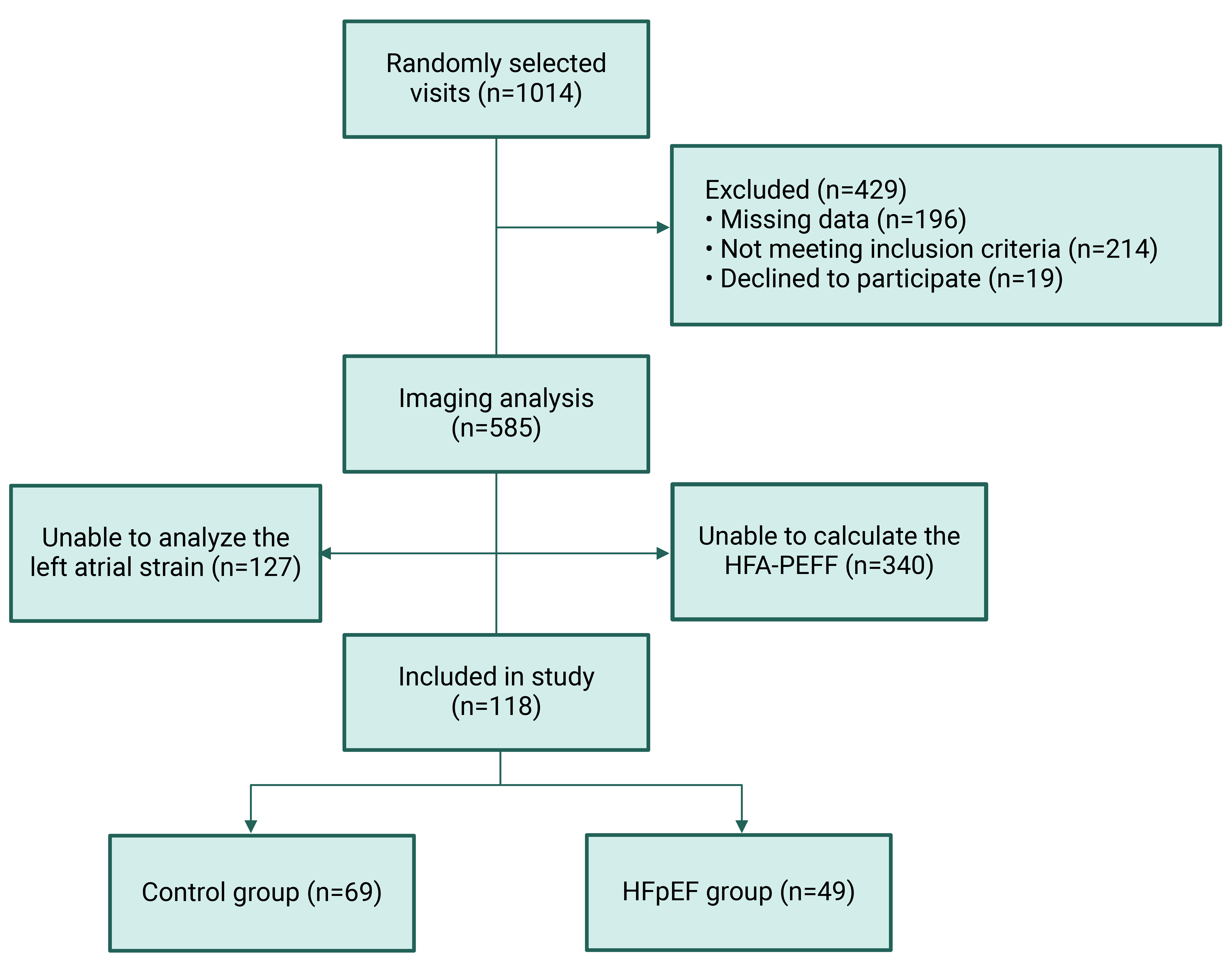

### Supplementary Figure 2

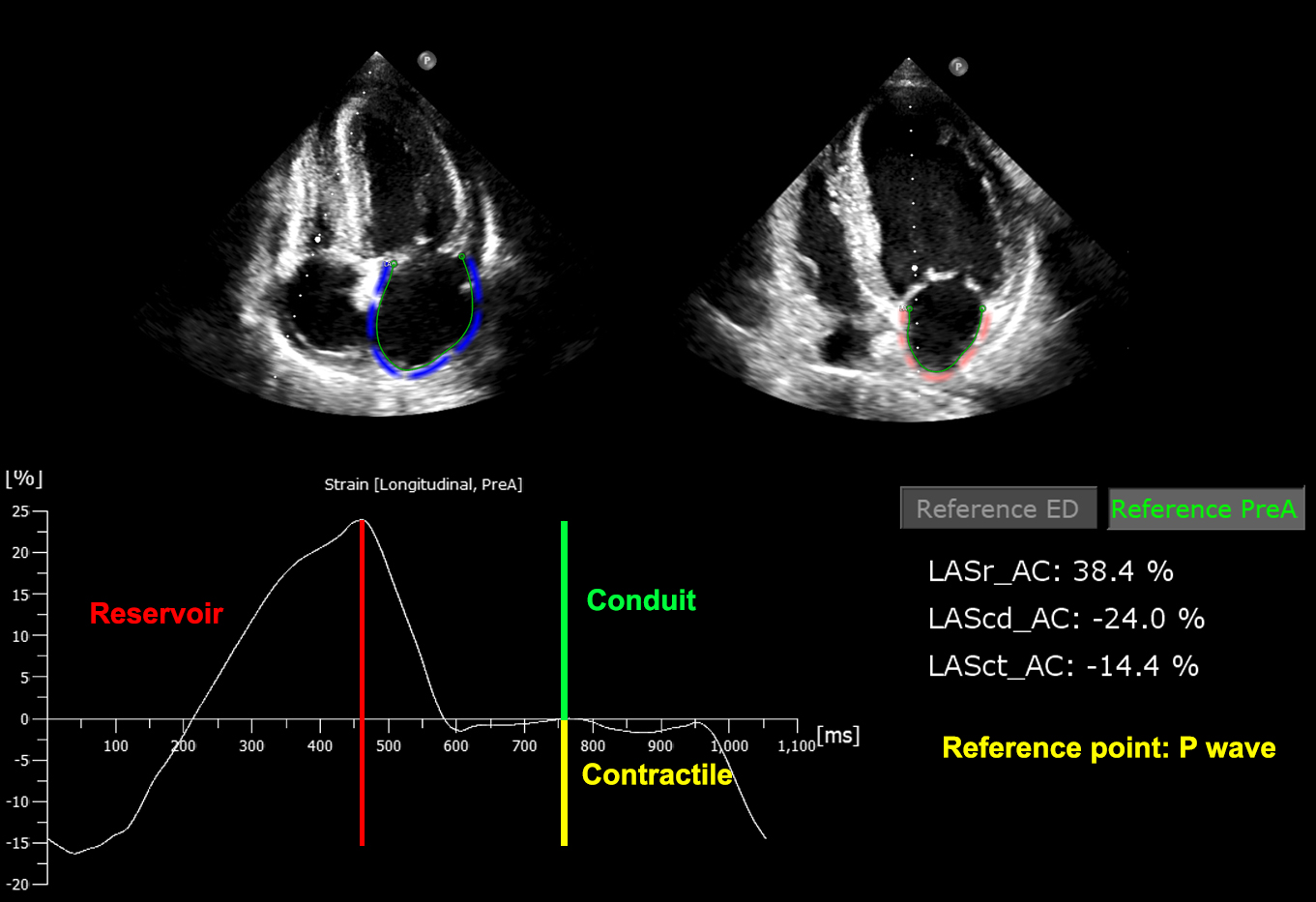
