## Supplementary Table 1 for "Comparing the value of left atrial strain and HFA-PEFF score in diagnosing heart failure with preserved ejection fraction: a cross-sectional study"

**Supplementary 1. Calculation of HFA-PEFF score**

| **HFA-PEFF score** | | | | |
| --- | --- | --- | --- | --- |
| **Parameter** | **Minor** | | **Major** | |
|  | **Value** | **Point** | **Value** | **Point** |
| **Functional** | Average E/e' 9 - 14  or GLS < 16% | 1 | Septal e' < 7 cm/s or lateral e' < 10 cm/s  or Average E/e' ≥ 15  or TR velocity > 280 cm/s  (PASP > 35 mmHg) | 2 |
| **Morphological** | LAVI 29 – 34 mL/m^2^ or LVMI ≥ 115/95 g/m^2^ (female/male) or  RWT > 0.42 or LV wall thickness ≥ 12 mm | 1 | LAVI > 34mL/m^2^ or LVMI ≥ 149/122 g/m^2^ (female/male) and RWT > 0.42 | 2 |
| **Biomarker (SR)** | NT-proBNP 125 - 220 pg/mL or BNP 35 - 80 pg/mL | 1 | NT-proBNP > 220 pg/mL or BNP > 80 pg/mL | 2 |
| **Biomarker (AF)** | NT-proBNP 365 - 660 pg/mL or BNP 105 - 240 pg/mL | 1 | NT-proBNP > 660 pg/mL or BNP > 240 pg/mL | 2 |

*Adapted from Heart Failure Association of the European Society of Cardiology*

Abbreviations: AF: atrial fibrillation; BNP: B-type natriuretic peptide; E: early transmitral flow velocity; e’: early diastolic mitral annular velocity; GLS: left ventricular global longitudinal strain; HFA-PEFF: Heart Failure Association-PEFF; LAVI: left atrial volume index; LV: left ventricle; LVMI: left ventricular mass index; NT-proBNP: N-terminal pro-B-type natriuretic peptide; PASP: pulmonary artery systolic pressure; RWT: relative wall thickness; SR: sinus rhythm; TRV: tricuspid regurgitation velocity.
